# Individual regional associations between Aβ-, tau- and neurodegeneration (ATN) with microglial activation in patients with primary and secondary tauopathies

**DOI:** 10.1101/2022.11.12.22282082

**Authors:** Anika Finze, Gloria Biechele, Boris-Stephan Rauchmann, Nicolai Franzmeier, Carla Palleis, Sabrina Katzdobler, Endy Weidinger, Selim Guersel, Sebastian Schuster, Stefanie Harris, Julia Schmitt, Leonie Beyer, Johannes Gnörich, Simon Lindner, Nathalie L. Albert, Christian Wetzel, Rainer Rupprecht, Axel Rominger, Adrian Danek, Lena Burow, Carolin Kurz, Maia Tato, Julia Utecht, Boris Papazov, Mirlind Zaganjori, Lena-Katharina Trappmann, Oliver Goldhardt, Timo Grimmer, Jan Haeckert, Daniel Janowitz, Katharina Buerger, Daniel Keeser, Sophia Stoecklein, Olaf Dietrich, Estrella Morenas-Rodriguez, Henryk Barthel, Osama Sabri, Peter Bartenstein, Mikael Simons, Christian Haass, Günter U. Höglinger, Johannes Levin, Robert Perneczky, Matthias Brendel

## Abstract

β-amyloid (Aβ) and tau aggregation as well as neuronal injury and atrophy (ATN) are the major hallmarks of Alzheimer’s disease (AD), and biomarkers for these hallmarks have been linked to neuroinflammation. However, the detailed regional associations of these biomarkers with microglial activation in individual patients remain to be elucidated.

We investigated a cohort of 55 patients with AD and primary tauopathies and 10 healthy controls that underwent TSPO-, Aβ-, tau-, and perfusion-surrogate-PET, as well as structural MRI. Z-score deviations for 246 brain regions were calculated and biomarker contributions of Aβ (A), tau (T), perfusion (N1) and gray matter atrophy (N2) to microglial activation (TSPO, I) were calculated for each individual subject. Individual ATN-related microglial activation was correlated with clinical performance and CSF soluble TREM2 (sTREM2) concentrations.

In typical and atypical AD, regional tau was stronger and more frequently associated with microglial activation when compared to regional Aβ (AD: β_T_ = 0.412±0.196 vs. β_A_ = 0.142±0.123, p < 0.001; AD-CBS: β_T_ = 0.385±0.176 vs. β_A_ = 0.131±0.186, p = 0.031). The strong association between regional tau and microglia reproduced well in primary tauopathies (β_T_ = 0.418±0.154). Stronger individual associations between tau and microglial activation were associated with poorer clinical performance. In patients with 4RT, sTREM2 levels showed a positive association with tau-related microglial activation.

Tau pathology has strong regional associations with microglial activation in primary and secondary tauopathies. An index of tau- and Aβ-associated microglia activation accounts for regional heterogeneity and allows for clinical and biomarker correlations with ATN-specific neuroinflammation.

## INTRODUCTION

Tauopathies account for the majority of neurodegenerative disorders. These, include the secondary 3/4-repeat tauopathy Alzheimer’s disease (AD) as the most prevalent form of dementia in societies with aging populations (1). Primary tauopathies, such as Pick’s disease, progressive supranuclear palsy (PSP) or corticobasal degeneration (CBD) are characterized by abnormal tau protein aggregation with three or four microtubule-binding-domains (2, 3). However, regardless of the distinct neuropathology, there is also overlap in clinical phenotypes, for example, in corticobasal syndrome (CBS), which can be characterized by 3/4-repeat AD-like tau or 4-repeat tau (4RT) aggregation (4).

The neuropathological cascade of AD is characterized by the triad of accumulation of extracellular β-amyloid (Aβ) plaques, fibrillary tau aggregates in neurons, and microgliosis and astrogliosis (5–9). In AD, this is conceptualized as the ATN sequence, where β-amyloidosis [A] is thought to precede and accelerate tau pathology [T] that leads to neurodegeneration [N] (10), now expanding towards an ATX(N) system, where X represents novel candidate biomarkers for additional pathophysiological mechanisms such as neuroimmune dysregulation, synaptic dysfunction or blood-brain-barrier alterations (11).

In AD and primary tauopathies, neuroinflammation represents an important mediator of protein aggregation and spread as well as disease progression, creating a positive feedback-loop in the disease cascade (12–15) and could serve as a valuable “X” parameter of ATX(N), then termed ATI(N). In this context, molecular imaging with 18kDa translocator protein (TSPO) positron-emission-tomography (TSPO-PET) for stratification and monitoring of neuroinflammation could become crucial for targeting and assessing response to immunomodulatory therapies and has spurned the development of a wide range of tracers for human studies (16–19). However, despite several studies with tau and TSPO tracers (20–22), ATI(N)-like schemes and interactions have only rarely been investigated in atypical AD or primary tauopathies.

A detailed understanding of the pathophysiological interplay between accumulation of misfolded proteins (i.e. Aβ and tau) and their contribution to neuroinflammation may provide deeper insight into mechanistic pathways (23). In recent years, several multi-tracer studies in cohorts composed mainly of patients along the typical AD continuum revealed discrepant findings. Some investigations attributed equal influence of Aβ and tau to microglial activation (24), while others supported the interaction of Aβ and microglia activation to set the pace for subsequent tau spread (12, 25). The effects of microglial activation on cognition are thought to be strongly mediated by gray matter atrophy (26), and activated microglia have been correlated with parietal atrophy post mortem (27). Thus, we aimed to investigate the interplay between ATI(N) biomarkers and microglial activation in a cohort composed of primary and secondary tauopathies, including atypical and typical AD. We analyzed the regional heterogeneity of ATI(N) biomarker alterations in tauopathies and we computed the partial contributions of ATN biomarkers to microglial activation at the individual patient level. Furthermore, we questioned the shape and magnitude of expurgated regional associations between aggregation of Aβ and tau with microglial activation. Finally, we correlated the individual Aβ- and tau-related microglial responses with clinical performance and cerebrospinal sTREM2 (soluble triggering receptor expressed on myeloid cells 2) levels.

## MATERIAL AND METHODS

### Study Design

We enrolled patients with typical AD, AD-CBS and 4RTs as well as healthy controls from ActiGliA, a prospective cohort study at Ludwig-Maximilians-University (LMU), approved by the local ethics committee (project-number 17-755; human PET analyses project-numbers 17-569, 19-022). Written informed consent was obtained from all participants in accordance with the Declaration of Helsinki. Requirements for inclusion were tau- and TSPO-PET, and T1w-MRI if available, within a time period of 18 months (mean time difference: 2.9 ± 4.4 months). Aβ-PET was tolerated up to 24 months prior to the remaining examinations (mean time difference: 6.7 ± 8.3 months). Tau-PET had to be conducted with dynamic emission recording to ensure availability of the early-phase as an “N” biomarker. Patients with a low affinity TSPO binding status were assigned to a separate “LAB” study group regardless of their clinical diagnosis.

Patients with dementia, mild cognitive impairment (MCI) or subjective memory decline (SCD) due to AD were diagnosed according to the diagnostic criteria of the National Institute on Aging (28). All patients with AD had a positive Aβ-PET scan and were allocated to the “typical AD” group. Clinical diagnosis of CBS was made as defined in the MDS-PSP criteria (29). All enrolled CBS patients also fulfilled the Armstrong criteria of probable or possible CBD-CBS (30). Only CBS patients with negative family history for Parkinson’s disease and AD were included. Positivity or negativity of the Aβ-PET scan defined the allocation into “AD-CBS” or “4RT” study groups.

Group level TSPO-PET data of this cohort was previously published for patients with AD, 4RTs and controls (31, 32). Group level tau-PET data of this cohorts was partially published for patients with AD, AD-CBS, 4RTs and controls (21). Group level data of early-phase imaging ([^18^F]flutemetamol and [^18^F]PI-2620) of parts of the cohort were previously published for AD-CBS and 4RTs (33, 34). All reported data are unpublished.

### PET imaging

*PET Data Acquisition, Reconstruction and Post-Processing:* For all PET procedures, including radiochemistry, acquisition and pre-processing, we used an established and standardized protocol (21, 31, 34, 35). Patients were scanned at the Department of Nuclear Medicine, LMU, using a Biograph 64 PET/CT scanner or a harmonized Biograph mCT (Siemens, Erlangen, Germany). In brief, [^18^F]GE-180 TSPO-PET recordings (average dose: 179 ± 13 MBq) with an emission window of 60-80 min after injection were obtained to measure microglial activation (31). [^18^F]flutemetamol Aβ-PET recordings (average dose: 185 ± 16 MBq) with an emission window of 90-110 min after injection were performed for assessment of fibrillar Aβ accumulation (35). Dynamic [^18^F]PI-2620 tau-PET (average dose: 188 ± 15 MBq) with emission recording 0-60 min after injection was performed to quantify tau aggregation. Static frames of the perfusion phase (0.5-2.5 min) (34) and the late phase (20-40 min) (36) were reconstructed.

*PET Image Analysis:* Several established processing pipelines (21, 31, 33, 34, 36) were executed to ensure reproducibility by different analysis methodology. We performed all PET data analyses using PMOD (V3.9; PMOD Technologies LLC; Zurich; Switzerland). The primary analysis used static emission recordings which were coregistered to the Montreal Neurology Institute (MNI) space using non-linear warping (16 iterations, frequency cutoff 25, transient input smoothing 8×8×8 mm³) to tracer-specific templates acquired in previous in-house studies (21, 31, 34, 35). Given the strong binding differences of positive and negative Aβ-PET images, we used positive (number of subjects in template: nt=15) and negative (n_t_=27) Aβ-PET templates after classification of the Aβ-status by a visual read by a single rater. A unified template was used for TSPO-PET (n_t_=11). Tau-PET images were coregistered to early (mixed [^18^F]PI-2620 and [^18^F]Flutemetamol; n_t_=15)(37) and late phase (n_t_=28) templates without distinction of tau-positivity.

Intensity normalization of all PET images was performed by calculation of standardized uptake value ratios (SUVr) using the cerebellum as an established pseudo-reference tissue for TSPO-PET (38). The cerebellum was selected as the best compromise of a unified pseudo-reference tissue since it was also validated for [^18^F]flutemetamol (39) and [^18^F]PI-2620. To account for tau-positive areas in the cerebellum and to avoid spill-over of adjacent extracerebral structures, the dentate nucleus as well as superior and posterior layers of the cerebellum were excluded. Using the Brainnetome atlas (40), the brain was parcellated into 210 cortical and 36 subcortical volume-of-interests (VOIs) and standardized-uptake-value-ratios (SUVr) were calculated for all four PET read-outs. Per subject, the standardized regional deviation of SUVr (z-score) was calculated versus previously established age- and sex-matched normal cohorts for [^18^F]flutemetamol (number of controls: n_c_=13, (33)), [^18^F]GE-180 (n_c_=13, (41)), early [^18^F]PI-2620 (n_c_=14, (42)) and late [^18^F]PI-2620 (n_c_=14, (4)).

### Structural MR imaging

Three-dimensional T1-weighted (T1w) MRI data were collected for the majority (60/72) of the cohort as reported previously (43). The data were coregistered into the MNI standard space using PNEURO tool (V3.9; PMOD Technologies LLC) and segmented into all 246 Brainnetome atlas VOIs. In this step, regional gray-matter-volumes (GMV), for analysis of possible brain atrophy, were collected. As described above, we used age- and sex-matched controls for calculation of standardized z-scores (n_c_=13).

### DNA extraction and SNP genotyping

Since the binding properties of second-generation TSPO ligands have been found to depend on genetic polymorphism of the TSPO gene (44, 45), all individuals underwent rs6971 single nucleotide polymorphism (SNP) genotyping and were classified as low, medium, or high affinity binder (LAB, MAB or HAB). For this purpose, whole-blood samples were sent to the Department of Psychiatry of the University Hospital Regensburg for genotyping. Genomic DNA was extracted from 4 ml of whole blood using a QIAamp DNA Blood Maxi kit (Qiagen, Hilden, Germany) according to the manufacturer’s protocol. DNA quality was assessed by optical absorbance and gel electrophoresis. Exon 4 of the TSPO gene and exon/intron junctions were amplified by PCR and sequenced using the Sanger method with the following primers: ex4-FAGTTGGGCAGTGGGACAG and ex4-R-CAGATCCTGCAGAGACGA. Sequencing data were analysed using SnapGene software (GSL Biotech; http://snapgene.com).

### sTREM2

sTREM2 concentrations were measured in all available CSF samples (4RT 14/26, AD-CBS 7/11, AD 10/18) by a modified assay based on the previously described sTREM2 ELISA using the MSD platform (46–48). This assay was designed to selectively detect sTREM2 coming from cleavage of the full-length protein. Prior to data analysis, the sTREM2 data were log-transformed.

### Clinical assessments

As a part of the ActiGliA protocol, patients underwent comprehensive clinical examinations. Some of these data were used to compare ATN-related microglial activation with clinical performance. In this work, the PSP-rating scale (PSPRS: [normal] x 0 – 100)(49), the Movement Disorder Society revised form of the Unified Parkinson’s Disease Rating Scale (UPDRS: x 0 – 100) (50) and the Geriatric Depression Scale (GDS: x 0 - 15) (51) were analyzed. In addition, the Montreal Cognitive Assessment scale (MoCA: 0 – 30 x) (52) and the Mini-Mental State examination (MMSE: 0 – 30 x) (53) were evaluated. For patients without MoCA but MMSE, the MMSE was converted to a comparable MoCA score (54). For simplicity, other clinical performance data collected by ActiGliA are not further mentioned.

### Statistics

For statistical calculations SPSS (V25; IBM; Ehningen; Germany) and R (V4.1.2; R Core Team; 2021; Vienna; Austria) were used. Unless declared otherwise, the significance level was set to 0.05. The day of Tau-PET examination was used to define the age of the patients.

#### Biomarker positivity classification

To assign patients to specific study groups, PET images were visually rated for Aβ-PET-positivity (single expert reader). In addition, an optimal cutoff based on a sensitivity / specificity tradeoff was defined for each of the 246 Brainnetome VOIs for each biomarker (excluding GMV) using single-region-ROC-analysis (cutoff: criterion of the point on the ROC curve closest to the point 0,1 (55, 56)). For semi-quantitative Aβ-PET we defined the threshold to best discriminate the AD groups (AD-CBS and typical AD) from 4RT and CTRL. For TSPO-PET, tau-PET, and perfusion, the optimal cutoff was determined to discriminate 4RT, AD-CBS and AD against CTRL. For every patient and biomarker we summed up the positivity of the five regions with highest area-under-curve (AUC) values and calculated another optimal cutoff via roc analysis to finally declare a patient as biomarker positive or negative.

#### Demographics, clinical performance and sTREM2

To test for group differences in demographic data, 1-way analysis of variance (ANOVA) were used for age, sTREM2 and MoCA. Sex and rs6971-SNP were subject to a chi-square (χ²) test. With significant ANOVA or χ²-test, multiple posthoc unpaired two-tailed Student’s *t*-tests were subsequently performed between all groups (unadjusted for multiple testing).

#### Multiregion analyses

A descriptive heatmap was created by calculating arithmetic means (x) within 24 composed Brainnetome regions (57) from the acquired z-scores and displayed in color scale per patient group and biomarker. Thereby, all x > 2 were set equal to 2 and all x < −2 were set equal to −2. For each of the 24 composite regions, the frequency of patients with peak z-scores (highest z-score along all 24 regions) was determined and visualized relative to the total amount of patients per group. These arithmetic means and biomarker peak distributions of the 24 brain regions were then tested for intercorrelations between those biomarkers (4RT, AD-CBS and AD pooled) using linear Pearson’s correlation (r).

#### Regression models

For each subject, a multiple linear regression model was defined to analyze the influence of the independent ATN variables (Aβ A, Tau T, Perfusion N1, GMV N2) on microglial activity (I), taking all 246 Brainnetome VOIs into account. The obtained standardized β-coefficients of ATN were visualized by a combined violin-box-plot. Within each diagnosis group, the β-coefficients of the four independent variables were tested for group differences using ANOVA with age and sex as covariates. Multiple post-hoc tests (dichotomous ANOVA, age and sex corrected) were performed if there was a significant group difference. False discovery rate (FDR, six tests) correction to decrease the risk of α-error-accumulation was applied. Furthermore, each p-value was Bonferroni adjusted for a total of five diagnostic groups.

As a supplementary analysis, similar to the methodology explained before, we ordered all β-values from the single-patient regression per biomarker and tested for group differences between the five diagnostic groups. Post-hoc analysis and correction for multiple testing were performed using FDR (ten tests) and Bonferroni (four biomarkers). To analyze the influence of ATN on microglial activity at group level, multivariate linear regression models (covariates: age, sex and subject-ID) were applied (including all 246 Brainnetome VOI z-scores of all patients per group). Partial regressions were analyzed and fitted values were used to classify a VOI as biomarker positive or negative. For ATN, the z-score cutoff for classification of positivity was determined at > 2 (AT) and < −2 (N). The z-score cutoff of I was set > 0, to ensure inclusion of regions with low-level inflammation above the average of controls. Proportions of I+A+ / I+T+ / I+N+ to the total amount of A+ / T+ / N+ VOIs were calculated.

#### Correlation and interaction analyses

Spearman’s rank correlation (r_s_) was used to analyze the relationship of clinical parameters and sTREM2 with AT-related microglial activation, in a cohort of composed 4RT, AD-CBS and AD patients. Correlation analyses were adjusted for sex, age, global-patient-TSPO-load (mean z-scores of 246 VOIs), and global-patient-AT-load (mean z-scores of 246 VOIs of the respective biomarker). For sTREM2, we separated the cohort into a group with AD-pathology (AD-CBS, AD) and a group with 4R-tauopathies. No correction for multiple testing due to the exploratory character was applied in this analyses. Subsequently, A and T were tested for interaction effects.

## RESULTS

### Demographics

The analyzed cohort consisted of 26 patients with 4RTs, 11 patients with AD-CBS, 18 patients with typical AD, 7 mixed LABs and 10 controls which received examinations with all ATN imaging biomarkers (**Table 1**). Age, sex and sTREM2 did not differ between subgroups of the cohort. A significant group difference for rs6971 SNP status was identified (4RT: MAB/HAB = 7/18, AD-CBS: MAB/HAB = 5/6, AD: MAB/HAB = 8/8, CTRL: MAB/HAB = 7/2, χ² = 8.575, p = 0.036). The MoCA scores of the controls were found to be significantly higher than those of 4RT, AD-CBS and AD. AD-CBS showed significantly lower MoCA scores than 4RT and AD (4RT: 22.8 ± 4.6, AD-CBS: 17.6 ± 6.6, AD: 22.4 ± 6.2, LAB: 22.0 ± 7.5, CTRL: 28.6 ± 1.6, F = 5.175, p = 0.001). Further demographic statistics are displayed in **Table 1**. Demographics of larger tracer specific control cohorts that were used for calculation of individual z-scores are reported in **Supplemental Table 1**.

**Table 1:**
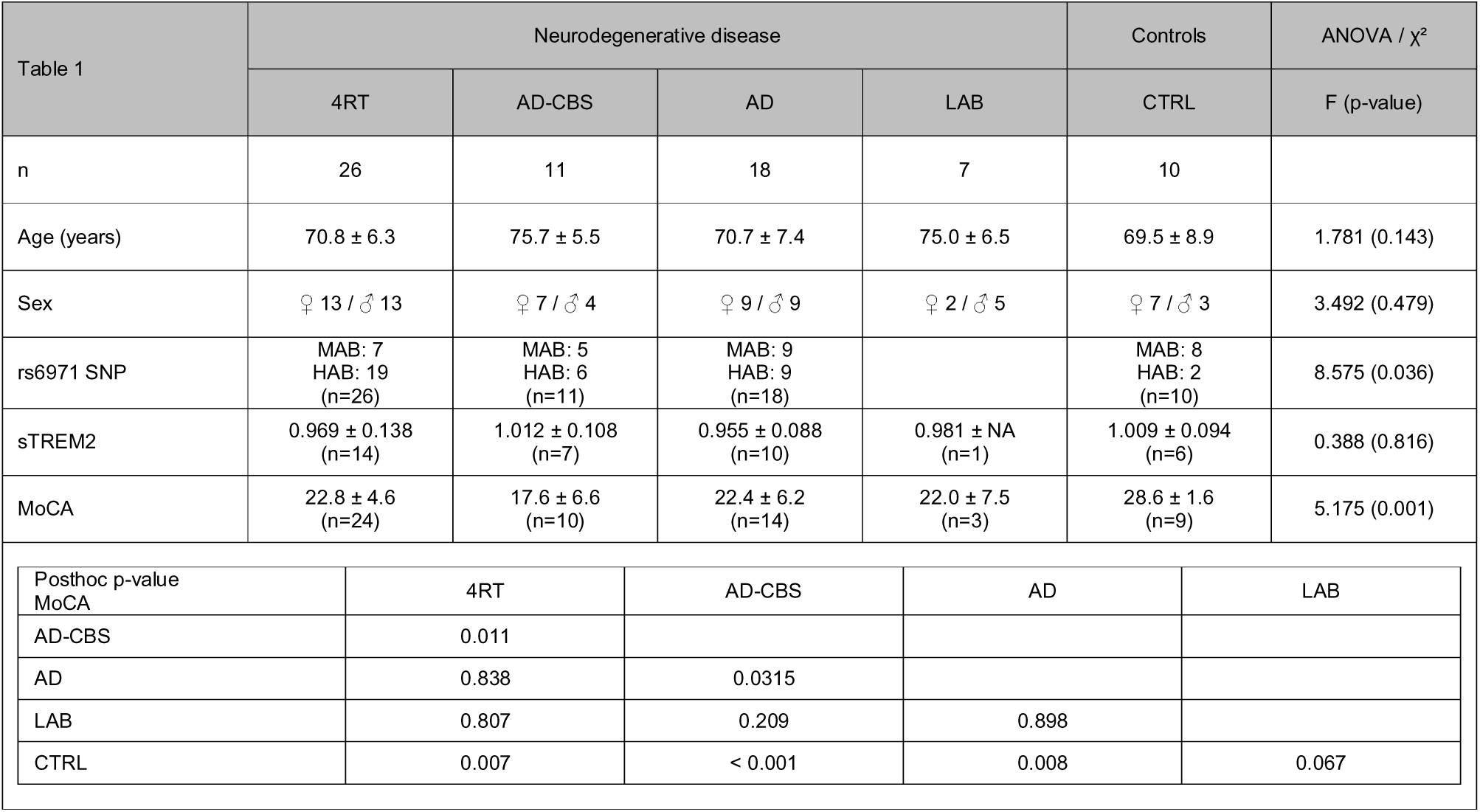
Demographics. Data are presented as mean ± standard deviation and number of frequency (n). Demographics were statistically tested by ANOVA or chi-square (χ²) test. With significant ANOVA group difference, multiple unpaired two-tailed Student’s t-tests were performed. Abbreviations: AD, Alzheimer’s disease; AD-CBS, corticobasal syndrome with AD-pathology; 4RT, four-repeat tauopathies; LAB, low affinity binder; MAB, medium affinity binder; HAB, high affinity binder; CTRL, healthy controls; ANOVA, analysis of variance; χ², chi-square; MoCA, Montreal-Cognitive-Assessment-Scale; MMSE, Mini-Mental State Examination; sTREM2, soluble triggering receptor expressed on myeloid cells 2.

### Strong regional heterogeneity of ATN biomarker alterations in primary and secondary tauopathies

First, we investigated regional alterations of microglial activity, Aβ, tau, perfusion and GMV in our cohort of primary (4RT) and secondary (typical AD, AD-CBS) tauopathies at the group level. Frequencies of overall biomarker positivity across subgroups are illustrated in **Figure 1A**. As expected from previous studies (4, 21) we found highest levels of Aβ-PET and tau-PET signals in cortical brain regions of patients with typical and atypical AD, whereas patients with 4RTs showed low Aβ-PET levels and subcortical predominance of tau accumulation (**Figure 1B**). TSPO-PET signal increases were abundantly observed in tau-, but not Aβ-PET positive brain regions (tau-TSPO: r = 0.312, p = 0.008 / Aβ-TSPO: r = 0.071, p = 0.552). Regional peak distribution of “N” biomarkers followed the individual patterns of tau- (tau-perfusion: r = 0.246, p = 0.037 / tau-atrophy: r = 0.270, p = 0.022), but not Aβ-PET (Aβ-perfusion: r = −0.023, p = 0.851 / Aβ −atrophy: r = 0.035, p = 0.771). TSPO-PET levels were distinctly higher in primary and secondary tauopathies with MAB and HAB status when compared to a mixed cohort (n=2 AD, n=4 4RT, n=1 healthy control) of low-affinity binders (**Figure 1C**).

**Figure 1:**
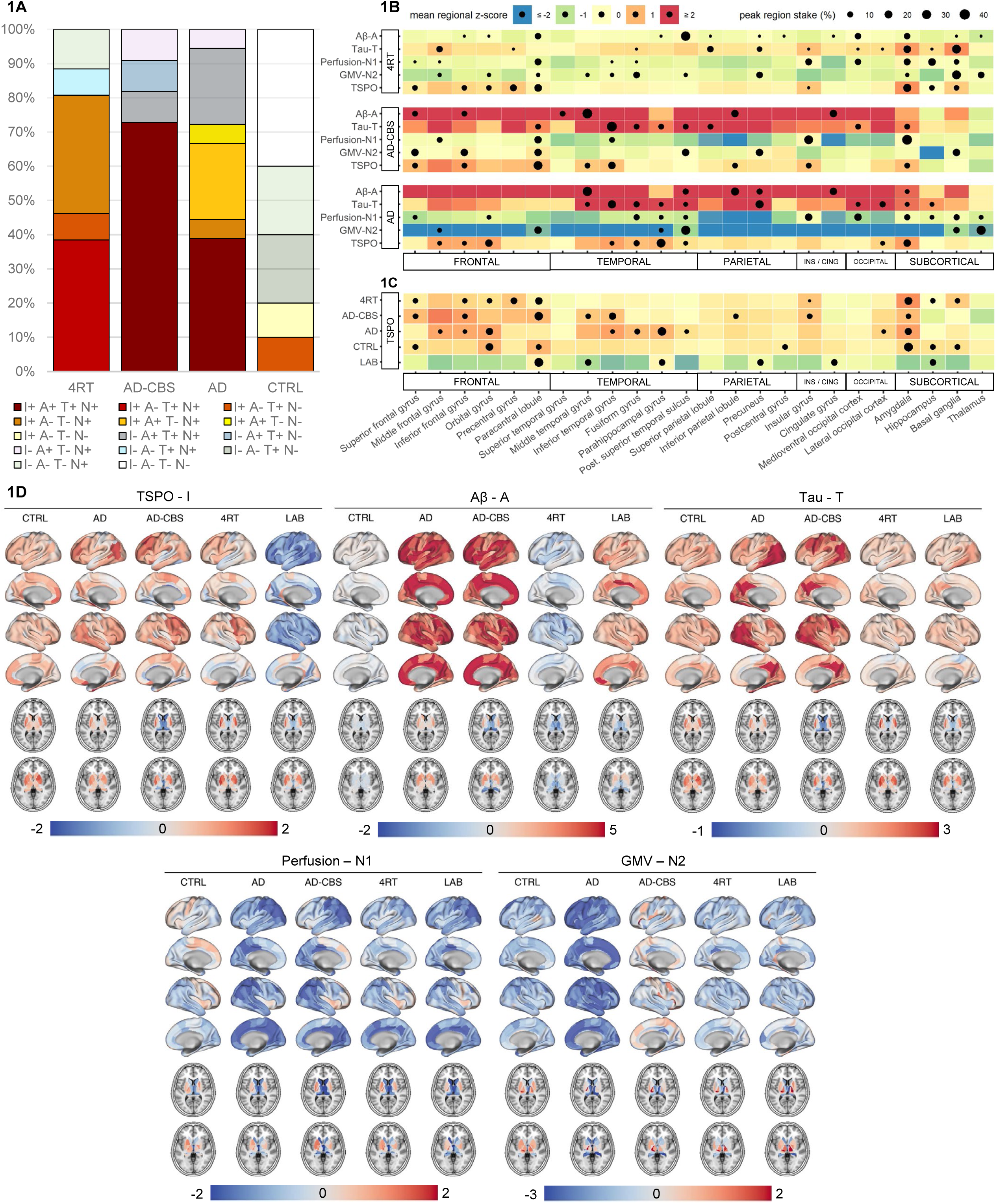
Determination of ATI(N) Biomarker Positivity and ATI(N) Biomarker Heterogeneity using a Multiregional Analysis. (A) Overall ATI(N) biomarker positivity of all included subjects for I(TSPO)-, A(Aβ)-, T(tau)- and N(perfusion, early tau)-PET at the group level. Single-region-ROC-analyses were applied to identify regions (n=5) that discriminate best among the study groups (maximum area-under-curve) for each biomarker. For each region an individual cutoff was set based on a sensitivity / specificity tradeoff and patients were classified as biomarker-positive or – negative based on the averaged regions that allowed the most optimal discrimination. Please note that AD also includes AD pathological change according to NIA-AA. (B) Heterogeneity of regional ATI(N) biomarker peak distribution. The heatmap was determined by calculating arithmetic means (x) within 24 composed brain regions (from - scores) and displayed in color scale. All x > 2 were set equal to 2 and all x < −2 were set equal to −2. The brain region with the highest z-score was determined for each study participant and the peak region frequency was plotted proportionally (%) to the number of patients of one group. (C) Comparison of TSPO-PET levels in patients with primary and secondary tauopathies and medium- and high-affinity TSPO binding status when compared to a mixed cohort (n=2 AD, n=4 4RT, n=1 healthy control) of individuals with low-affinity TSPO binding status using the same methodological approach as in (B). (D) Regional ATI(N) biomarker PET signal alterations at the group level. Within each study group, the mean z-score of each Brainnetome region was calculated and plotted as surface projections (lateral and medial view) and axial slices of the basal ganglia. *Abbreviations*: AD, Alzheimer’s disease; AD-CBS, corticobasal syndrome with AD-pathology; 4RT, four-repeat tauopathies; LAB, low affinity binder; CTRL, healthy controls; TSPO, 18-kDa translocator protein; Aβ, β-amyloid; GMV, gray-matter-volume; PET, positron-emissions-tomography; ROC, receiver-operating-characteristic

Importantly, we found a scattered distribution of individual ATN peak regions within subgroups of primary and secondary tauopathy subgroups (Figure 1D). Highest Aβ-PET z-scores in individual patients with typical AD were observed in temporal (44.4%) and parietal lobe (33.3%), as well as cingulate gyrus (16.7%) and amygdala (5.6%), whereas individual patients with AD-CBS had highest Aβ-PET z-scores in the middle and superior temporal gyrus (45.5%), parietal lobe (18.2%), cingulate (18.2%) and frontal cortices (18.2%). As expected from previous work (4), highest individual tau-PET alterations in patients with 4RTs were observed in the basal ganglia (26.9%), but heterogeneity of peak signal alterations was observed with involvement of limbic regions (23.1%), parietal (19.2%), frontal (15.4%) and temporal (3.8%) cortices. In line with previous pattern classification (58), we found various peak regions of individual tau-PET not only in AD-CBS but also in typical AD, with predominance of temporal regions (50.0%) but peaks in the precuneus (22.2%), occipital cortices (16.7%) as well as limbic regions (11.1%).

### Regional Tau-PET is closely associated with the individual microglia response in primary and secondary tauopathies

Given the strong regional heterogeneity of Aβ and tau accumulation in primary and secondary tauopathies, we applied a multi-regional regression model to investigate the detailed associations of ATN biomarker alterations with microglia activation at the individual patient level (**Figure 2A**). In 4RTs, Tau-PET contributed significantly to the regional explanation of TSPO-PET (β_T_ = 0.418 ± 0.154 / p < 0.001 in 88% of individuals), whereas Aβ-PET did not (β_A_ = −0.030 ± 0.124 / p < 0.001 in 15% of individuals; **Figure 2B/C**).

**Figure 2:**
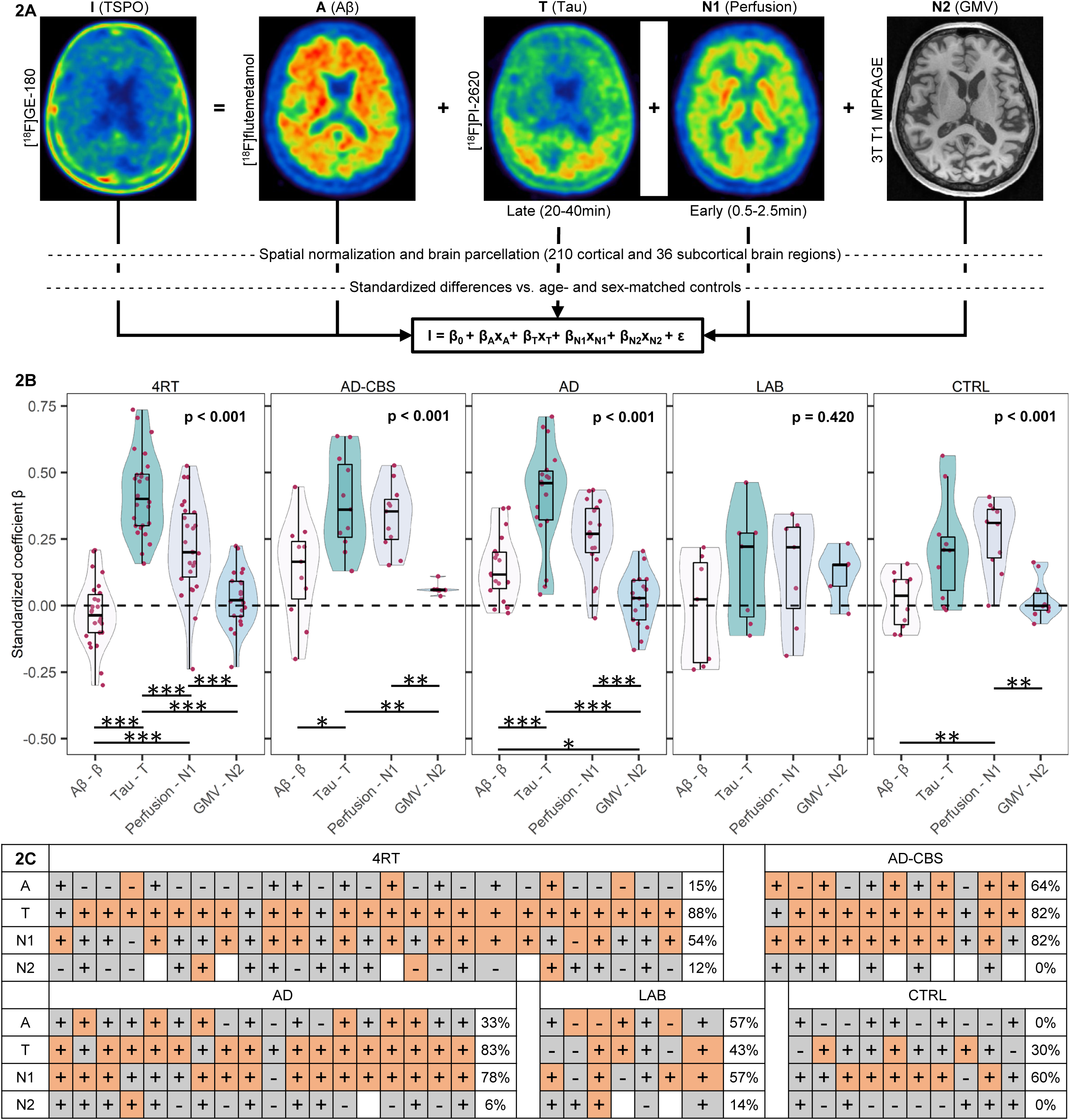
Multiregional Regression Analysis to Determine the Association of ATN Biomarkers with Microglial Activation at the Individual Patient Level. (**A**) Methodological work flow: PET and MRI data were spatial normalized to tracer specific templates acquired in previous in house studies (Montreal Neurology Institute (MNI) space) and parcellated into 210 cortical and 36 subcortical brain regions. PET-tracer uptake and gray-matter-volumes (GMV, from MRI) were transformed into z-scores with age- and sex-matched controls. A multiple linear regression model was performed (I = β_0_ + β_A_x_A_ + β_T_x_T_ + β_N1_x_N1_ + β_N2_x_N2_ + ε) at the single-patient-level and standardized coefficients (β) were derived for each ATN biomarker. (**B**) β-coefficients were visualized as a combined violin-box-plot and show the association of ATN biomarkers with microglia activation, each dot representing a single subject. Within each diagnosis group, the β-coefficients of the four independent variables were tested for group differences using ANOVA with age and sex as covariates. Multiple posthoc tests (dichotomous ANOVA, age and sex corrected) were performed if there was a significant group difference. Combined false discovery rate (FDR, six p-values) and Bonferroni (five p-values) correction were applied to decrease risk of α-error-accumulation. * p < 0.05, ** p < 0.01, *** p < 0.001. (**C**) Heatmap indicates the significance of ATN biomarker contribution to microglia activation resulting from the regression analysis in different diagnostic subgroups. Orange: p < 0.001. Gray: p ≥ 0.001. The percentage was calculated as the proportion of significant regression models of patients, relative to the total number of patients in the diagnostic group. Plus and minus signs were used to indicate the positivity or negativity of the associated β-coefficients. Abbreviations: AD, Alzheimer’s disease; AD-CBS, corticobasal syndrome with AD-pathology; 4RT, four-repeat tauopathies; LAB, low affinity binder; CTRL, healthy controls; TSPO, 18-kDa translocator protein; Aβ, β-amyloid; GMV, gray-matter-volume; PET, positron-emissions-tomography; MRI, magnetic-resonance-imaging; MNI, Montreal Neurology Institute; ANOVA, analysis of variance; FDR, false discovery rate.

Perfusion (β_N1_ = 0.218 ± 0.176 / p < 0.001 in 54% of individuals) and GMVs (β_N2_ = 0.026 ± 0.108 / p < 0.001 in 12% of individuals) were associated with regional TSPO-PET in individuals with 4RTs but did not exceed controls at the group level (**Supplemental Figure 1**). The positive character of these regional associations suggested an unspecific coupling of regional tracer signals by perfusion and atrophy. Within the 4RT subgroup, explanation of TSPO-PET alterations by tau-PET distinctly exceeded the explanation by “A” and “N” biomarkers (**Figure 2B**).

In patients with AD-CBS and typical AD, we observed significant explanation of regional TSPO-PET by Aβ-PET (AD-CBS: β_A_ = 0.131 ± 0.186 / p < 0.001 in 64% of individuals; AD: β_A_ = 0.142 ± 0.123 / p < 0.001 in 33% of individuals) and tau-PET (AD-CBS: β_T_ = 0.385 ± 0.176 / p < 0.001 in 82% of individuals; AD: β_T_ = 0.412 ± 0.196 / p < 0.001 in 83% individuals; **Figure 2B/C**), but explanation by perfusion and GMVs did again not exceed the level of controls (**Supplemental Figure 1**). Tau-PET indicated the strongest regional relations with TSPO-PET in both AD groups among all ATN biomarkers (**Figure 2B**).

In the subgroup of mixed LABs, we did not observe a significant explanation of the TSPO-PET signal by ATN biomarkers (p = 0.420). Associations between tau-PET and Aβ-PET with TSPO-PET were obliterated in individual cases with LAB status (**Supplemental Figure 2**). Noteworthy, the individual positive associations between perfusion and TSPO-PET were still present for cases with LAB status, pinpointing the unspecific regional association between perfusion and TSPO uptake patterns when the tracer cannot bind the target.

### Incremental tau aggregation is associated with high regional activation of microglia

To test for regional associations between ATN biomarkers and microglia activation at the disease level, we applied a partial regression model to study the specific association of single ATN parameters with TSPO-PET, controlled for all other ATN biomarkers, age, sex and subject-ID.

When pooling VOI-specific data across 4RT subjects (26 patients with 246 Brainnetome VOIs), 6396 analyzed brain regions revealed high proportions of TSPO-positive (I+) regions among T+ (82%) but a low proportion of TSPO-positive regions among A+ (48%) and N+ (27%) regions (**Figure 3A**). There was a weak partial association between regional Aβ(β= 0.030, p = 0.011) with TSPO but a strong partial association between regional Tau and TSPO (β = 0.381, p < 0.001; **Figure 3A**). The partial association between perfusion and TSPO (β = 0.270, p < 0.001) was driven by lower TSPO-PET signals in regions with strong decreases in perfusion (**Figure 3A**).

**Figure 3:**
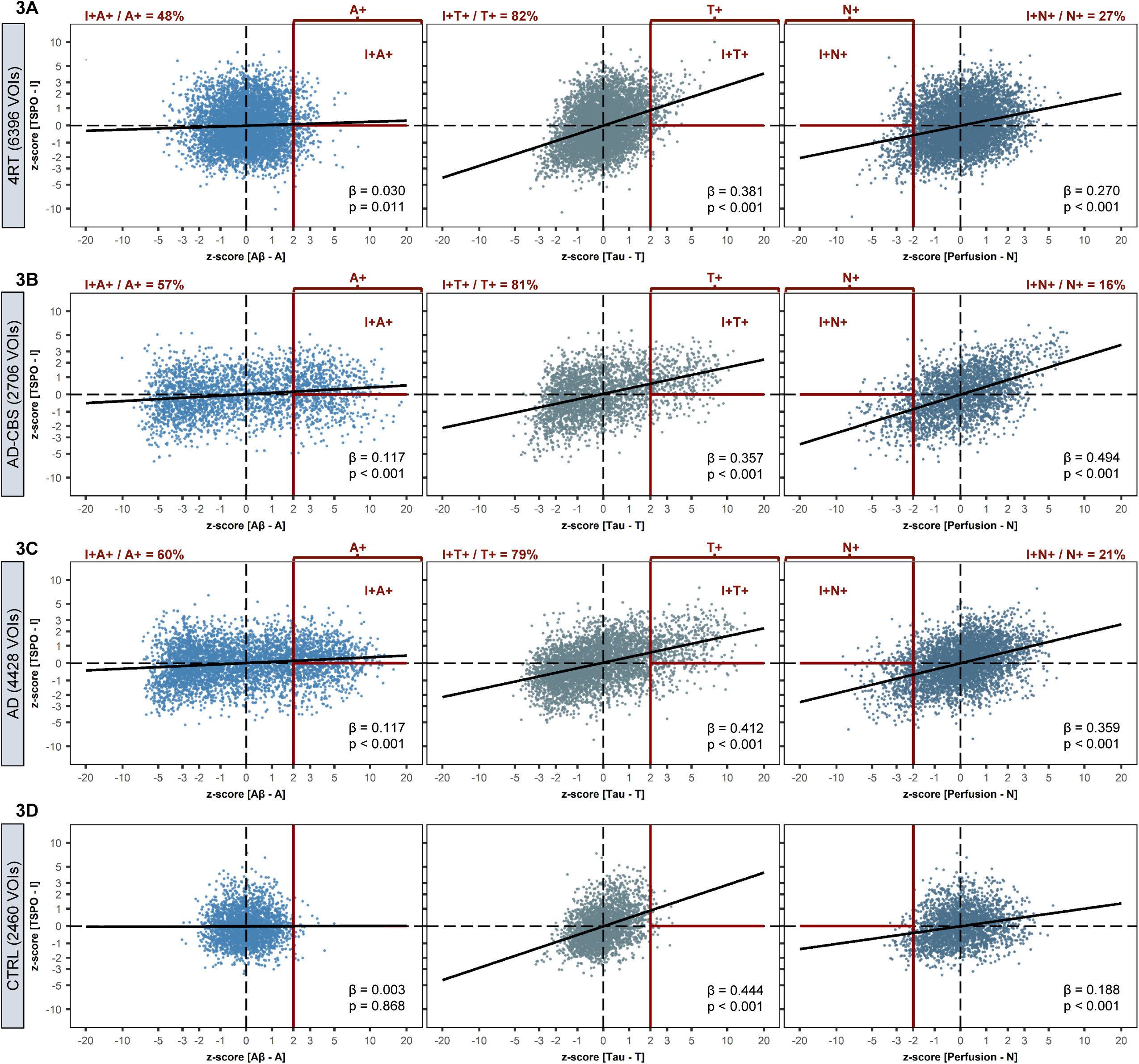
Multiregional Regression Analysis to Determine the Association of ATN Biomarkers with Microglial Activation at the Group Level. A multiple linear regression model was performed (I = β_0_ + β_A_x_A_ + β_T_x_T_ + β_N_x_N_ + age + sex + subject-ID + ε) at group-level for (A) 4RT, (B) AD-CBS, (C) AD and (D) CTRL using z-scores of all single regions of all individuals combined. The standardized coefficient (β) and the associated p-value were calculated. Fitted values of the partial regressions were used to classify a VOI as biomarker positive or negative. For ATN, the z-score cutoff for classification of positivity was determined at > 2 (AT) and < −2 (N). The z-score cutoff of I was set > 0. Proportions of I+A+ / I+T+ / I+N+ to the total amount of A+ / T+ / N+ VOIs were calculated. Abbreviations: AD, Alzheimer’s disease; AD-CBS, corticobasal syndrome with AD-pathology; 4RT, four-repeat tauopathies; CTRL, healthy controls; TSPO, 18-kDa translocator protein; Aβ, β-amyloid.

Both AD cohorts (2706 analyzed regions of AD-CBS and 4428 analyzed regions of typical AD) revealed similar partial associations between regional ATN biomarker alterations and TSPO-PET. There was a weak but significant regional association between Aβ and TSPO in patients with AD-CBS (β = 0.117, p < 0.001) and typical AD (β = 0.117, p < 0.001; **Figure 3B/C**). Regional tau again appeared as the strongest driver of TSPO-PET signals in AD-CBS (β = 0.357, p < 0.001) and typical AD (β = 0.412, p < 0.001) with high proportions of regional TSPO-PET positivity in regions with significant tau accumulation (AD-CBS: 81%, typical AD: 79%; **Figure 3B/C**). Perfusion indicated a significant association with TSPO-PET signals for both AD subgroups (**Figure 3B/C**). Controls validated the applied thresholds of ATN positivity and indicated only few A+, T+ and N+ regions (**Figure 3D**).

### Associations of Aβ- and tau-associated microglial response with clinical performance and sTREM2

Finally we tested if individual associations between Aβ and tau with microglial activation were correlated with indices of clinical severity and sTREM2. The rationale was to investigate if specific “A”or “T” associated microglial activation can be linked to improved or deteriorated clinical performance. Tau associated microglial activation revealed a general association with worse clinical performance (**Figure 4**). Patients indicated more depressive symptoms (r_s_ = 0.460, p = 0.011) and worse results on the UPDRS scale (r_s_ = 0.503, p = 0.006), when patients had high associations between regional tau and microglial activation. We observed a significant interaction effect between Aβ and tau associated microglial activation for UPDRS (T = 2.401, p = 0.019) and GDS (T = 3.581, p < 0.001), suggesting that microglial response to different protein aggregation may have opposite effects on brain function and clinical severity. There were no correlations, nor interaction effects for MoCA or PSPRS. The AD-group showed a significant interaction effect when Aβ-associated TSPO-PET signals and tau-associated TSPO-PET signals were correlated with sTREM2 (T = 2.732, p = 0.011; **Figure 5**). This indicated that high Aβ-TSPO associations but low tau-TSPO associations are linked with sTREM2 increases at the individual patient level. Contrary, patients with 4RT showed high sTREM2 levels when regional TSPO-PET was associated with tau-PET (r_s_ = 0.705, p = 0.034), may indicating different trajectories of microglia phenotypes in the course of primary and secondary tauopathies.

**Figure 4:**
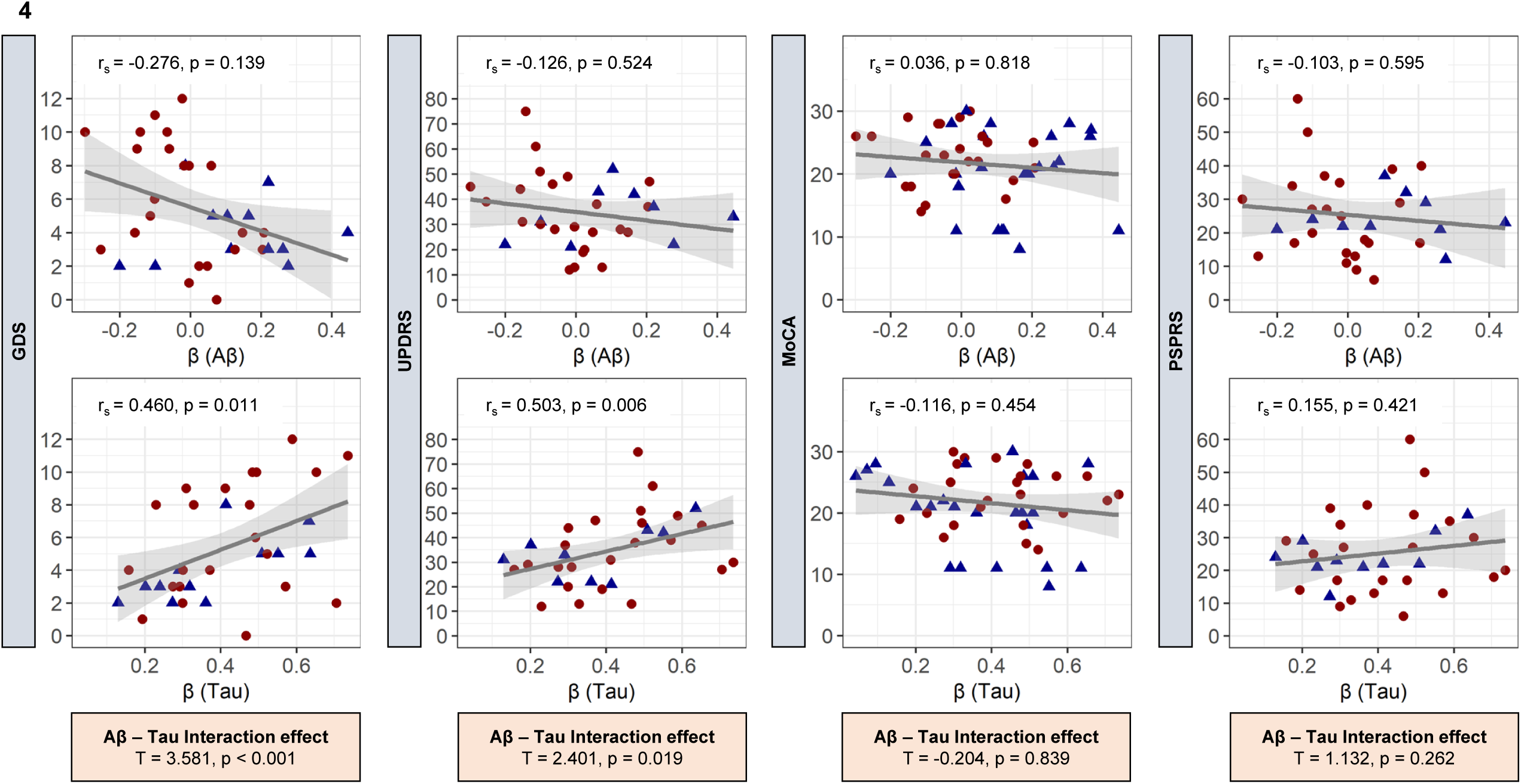
Correlation of AT Associated Microglial Activation with Clinical Performance. Standardized coefficients β (single patient regression), to detect Aβ (A)- and tau (T)-associated microglial activation, were correlated with clinical performance scores (Spearman’s correlation coefficient, r_s_). For statistical calculations 4RT, AD-CBS and AD were pooled. 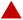 = 4RT, 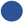 = AD-CBS + AD. Correlation analyses were adjusted for sex, age, global-patient-TSPO-load (mean z-scores of 246 VOIs), and global-patient-AT-load (mean z-scores of 246 VOIs of the respective biomarker). No correction for multiple testing was applied. Subsequently, A and T were tested for interaction effects. Abbreviations: AD, Alzheimer’s disease; AD-CBS, corticobasal syndrome with AD-pathology; 4RT, four-repeat tauopathies; LAB, low affinity binder; CTRL, healthy controls; TSPO, 18-kDa translocator protein; Aβ, β-amyloid; GMV, gray-matter-volume; MoCA, Montreal-Cognitive-Assessment-Scale; MMSE, Mini-Mental State Examination; UPDRS, Unified-Parkinson’s-Disease-Rating-Scale; PSPRS, Progressive-Supranuclear-Palsy-Rating-Scale; GDS, Geriatric-Depression-Scale.

**Figure 5:**
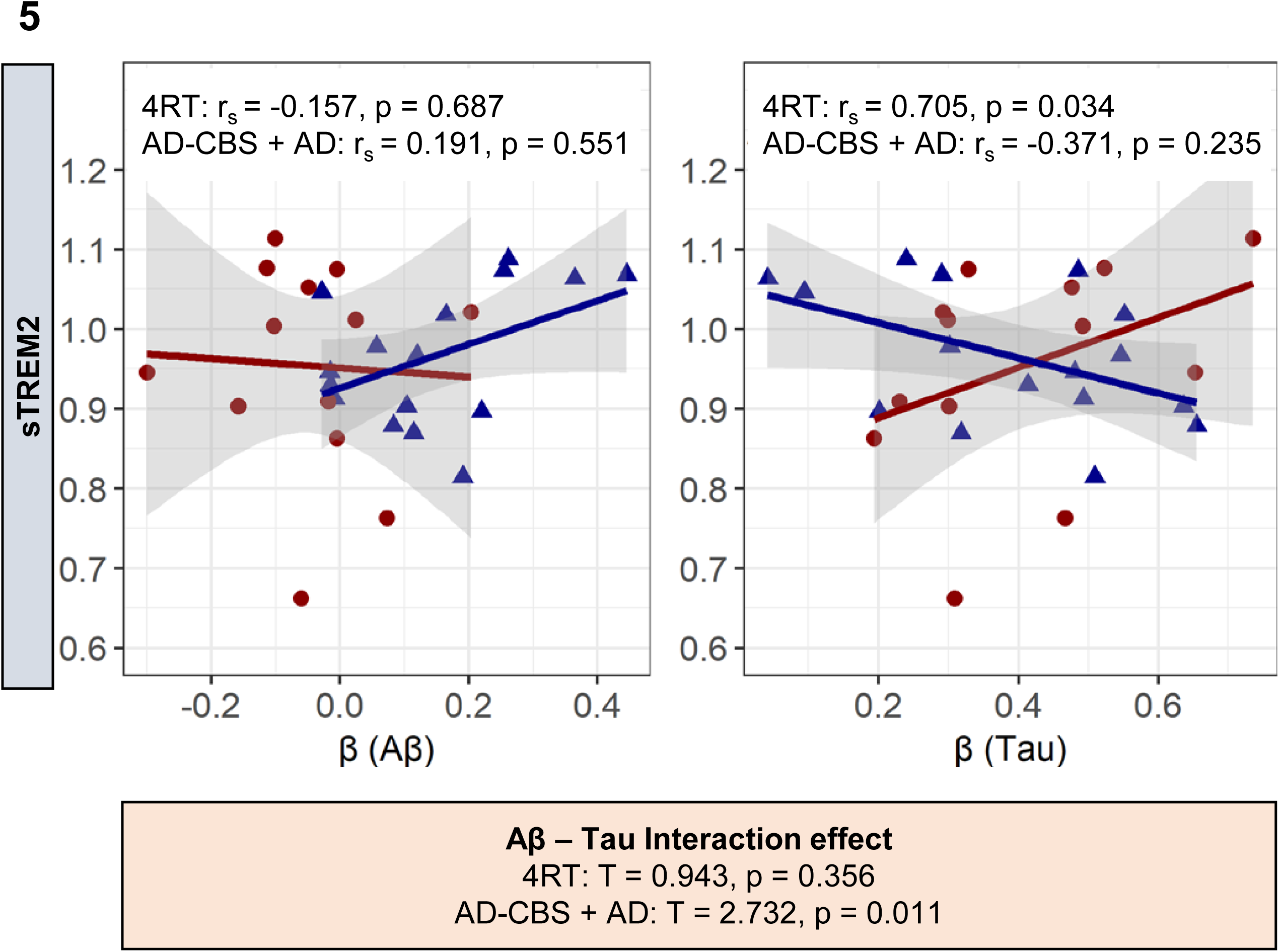
Correlation of ATN Associated Microglial Activation with sTREM2. Standardized coefficients β (single patient regression), to detect Aβ (A)- and tau (T)-associated microglial activation, were correlated with sTREM2 data (Spearman’s correlation coefficient, r_s_). The statistical analysis was each calculated for the combined AD group (AD-CBS + AD), and the 4RT group. 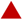= 4RT, 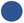 = AD-CBS + AD. Correlation analyses were adjusted for sex, age, global-patient-TSPO-load (mean z-scores of 246 VOIs), and global-patient-AT-load (mean z-scores of 246 VOIs of the respective biomarker).No correction for multiple testing was applied. Subsequently, A and T were tested for interaction effects. Abbreviations: AD, Alzheimer’s disease; AD-CBS, corticobasal syndrome with AD-pathology; 4RT, four-repeat tauopathies; LAB, low affinity binder; CTRL, healthy controls; TSPO, 18-kDa translocator protein; Aβ, β-amyloid; GMV, gray-matter-volume; sTREM2, soluble triggering receptor expressed on myeloid cells 2; PSPRS, Progressive-Supranuclear-Palsy-Rating-Scale; MMSE, Mini-Mental State Examination.

## DISCUSSION

We provide the first study that uses regional resolution of ATI(N) PET and grey matter volumes in a combined cohort of primary and secondary tauopathies to investigate the detailed associations between accumulation of misfolded proteins, perfusion and volume changes with microglial activation. We find distinct heterogeneity of regional distribution patterns of ATI(N) biomarkers among patients with tauopathies. Regional accumulation of tau was the strongest predictor of microglial activation in individual patients with primary and secondary tauopathies, whereas Aβ, perfusion and gray matter volumes indicated only weaker associations with regional TSPO-PET. Individual associations between ATN biomarkers and microglial activation were abrogated in low-affinity binders, demonstrating *in vivo* specificity of [^18^F]GE-180 to TSPO. Finally, we obtained preliminary evidence of worse clinical performance when microglial activation was associated with tau at the individual patient level.

The major finding of our study revealed that, among all ATN biomarkers, aggregated tau has the closest link with colocalized microglia activation in primary tauopathies and AD. Thus, our study confirmed joint accumulation of tau and microglial activation in typical AD (12) and adds evidence of similar regional trajectories in 4RTs and atypical AD. Several clinical and preclinical investigations indicated that both Aβ and tau are closely related to PET and histological measurements of increased microglial activity (12, 15, 24, 59–61). However, the detailed interplay between ATN biomarkers and neuroinflammation was discussed controversially. Even at the stage of single ATN biomarkers, distinct subtypes of microglia activation have been shown to occur in response to Aβ accumulation, depending on disease stage (62): a protective neuroinflammatory response that could be beneficial due to phagocytosis of neurotoxic Aβ species, and a detrimental one with negative effects that leads to inhibition of synaptic functioning and increased cell death (62, 63). Furthermore, atrophy was found to mediate effects of both tau pathology and neuroinflammation on cognition in AD (26). We hypothesized that regression models of multiple biomarkers in different brain regions could deepen the understanding of the ATI(N) biomarker interplay with I as an index of microglial activation. The application of this approach at the individual patient level enabled us to directly compare the partial influence of ATN biomarkers and we clearly observed a stronger association between regional aggregation of tau with microglial activation than did Aβ advanced stages of Aβ deposits. This finding could be related to exhausted microglia at accumulation (64). Patients with typical and atypical AD showed a significant influence of regional Aβ on microglial activation but this association was not found in 4RTs. This is of particularly interest since subthreshold levels of Aβ occur in 4RTs (65) but do not have a major impact on neuroinflammation according to our data.

With regards to therapy monitoring, it is of tremendous interest to understand associations between ATN biomarkers and neuroinflammation during the progressive aggregation of misfolded proteins in the brain. Thus, we also applied a model comprising all individual regional ATN biomarker data within groups of patients with 4RTs, typical AD and atypical AD, again exploring the associations of the individual ATN biomarker adjusted for the remaining indices. Importantly, we found that, in a cross-sectional design, microglial activation still continuous to rise coupled with increasing tau pathology at stages with very high tau accumulation in primary and secondary tauopathies (**Figure 3**). Contrary, TSPO-PET associations with regional Aβ aggregation tended to peak or plateau at stages of moderate Aβ abundance. This observation fits to a recent study, suggesting that microglial activation correlates strongly with tau aggregation in established AD and with Aβ deposition in MCI (24). Furthermore, two other studies indicated specific associations between Aβ and neuroinflammation at early stages of Aβ pathology (66, 67). Thus, targeting tau associated microglial activation could still be beneficial at later stages of disease, whereas modifying Aβ associated microglial activation may be only beneficial at early disease stages (48). Understanding differences of microglia phenotypes associated either with Aβ or tau aggregation will likely have a major impact for selection of appropriate immunomodulatory strategies.

The relevance of regression models at the individual patient level was pinpointed by the observation of distinct heterogeneity of regional ATN biomarker alterations in patients with 4RTs and AD. In line, four distinct trajectories of tau deposition were recently identified in typical AD, which implies that pathology originates and spreads through distinct networks in the different subtypes (58, 65). Distinct regional patterns of neurodegeneration were similarly reported for 4RTs (68). This heterogeneity implies that conservative regional assessments at the group level limit the possibility to capture the full picture of ATN biomarker alterations because effects in single regions are diminishes by differing regional peaks of pathology.

An exploratory analysis of our study suggested that strong regional associations between tau and microglial activation indicated worse clinical performance at a cross-sectional level of analysis in patients with primary and secondary tauopathies. This finding is in line with improved cognition in tauopathy models with NLRP3 inflammasome knock-out (69). In contrast, the individual regional association of microglial activation with Aβ showed a more beneficial effect on clinical performance indices in patients with AD. Supporting this claim, levels of sTREM2 were specifically associated with Aβ related but not tau related microglial activation in our cohort, while sTREM2 was reported to be associated with reduced cognitive decline in AD (70). In this regard, TREM2 may slow AD progression and reduce tau-driven neurodegeneration by restricting the degree to which β-amyloid facilitates the spreading of pathogenic tau (71). Our observations also had similarities to serial preclinical studies in mouse models, where microglial activation was correlated with worse outcome parameters in the P301S tau model (60), but with improved spatial learning performance in PS2APP (72, 73) and *App^NL-G-F^* (17) Aβ models. A clinical study in patients with AD also suggested that anterior temporal [^11^C]PK11195 binding, a region with abundant tau but low Aβ deposition is associated with progressive cognitive decline (74). In this regard, our individual index of tau- and Aβ-associated microglia activation could serve to account for regional heterogeneity and standardized quantification of the multidimensional neuroinflammatory response. This may enable stratification for immunomodulatory therapies by identification of patients with high risk of future clinical deterioration (74).

The characteristics of the used molecular imaging biomarkers for assessment of ATI(N) need to be considered for interpretation of our results. A strength of this study is given by the use of the novel tau-PET tracer [^18^F]PI-2620 which proved absent off-target binding to monoamine oxidases (75), high affinity to 3/4R-tau in AD (76) and also high affinity to recombinant 4R tau fibrils and PSP brain homogenate (75). Colocalization of [^18^F]PI-2620 binding with 4R tau in micro-autoradiography (75) and quantitative agreement of autoradiography and immunohistochemistry (65, 77) as well as *in vivo* discrimination of PSP (21) and CBS (4) from controls highlight the potential of this radioligand. Furthermore, early-phase [^18^F]PI-2620 PET indicated feasibility for perfusion surrogate imaging in patients with AD and 4RTs (34, 78), which was applied a “N” biomarker in the current study. [^18^F]flutemetamol is an FDA and EMA approved Aβ-PET tracer and characterized by fast brain uptake, fast clearance, and excellent binding properties to Aβ deposits *in vitro* and *in vivo* (79). TSPO-PET represents a sensitive biomarker for monitoring of immunomodulation and can act as a surrogate for activated microglia. Several TSPO radioligands allow monitoring neuroinflammation in AD and 4R-tauopathies (18) and we were able to show that [^18^F]GE-180 recapitulates expected topology of microglial activation in 4RTs (22) and AD (43). However, [^18^F]GE-180 has recently been criticized for limited uptake across the blood-brain-barrier (22, 80), since blood-brain integrity can be disturbed in neurodegenerative disorders with a potential impact on tracer kinetics (81, 82). We circumvented blood-brain-barrier related perfusion alterations and interrogated them as a potential cofounder by inclusion of a perfusion surrogate as an “N” biomarker into our regression model. With this approach, we were able to show that perfusion has a substantial impact on [^18^F]GE-180 binding, but our model also allowed us to determine the specific perfusion-adjusted impact of tau and Aβ pathology on TSPO-PET signal alterations. More importantly, we were able to show that associations between regional tau and Aβ accumulation with microglial activity were lost in low-affinity-binders. Furthermore, regional TSPO-PET signals were distinctly lower in participants with low-affinity binding status when compared to medium and high affinity binders. Thus, our data provide evidence on the specificity of the [^18^F]GE-180 TSPO-PET signal in patients with neurodegenerative diseases.

Limitations of the study include that we cannot draw detailed conclusions about the cell type specificity of the TSPO-PET signal to activated microglia, since we did not measure associations with reactive astrocytosis by glial fibrillary acidic protein or a PET radioligand targeting reactive astrocytes. Furthermore, our study enrolled patients based on state-of-the-art clinical diagnosis but was not enriched by neuropathological confirmation. Thus, although the characterization of our patients was based on several biomarkers and detailed clinical work-up, misdiagnosed patients need to be considered as potential confounders in this study population.

## CONCLUSION

A multi-regional regression model of PET and grey matter volume can serve for personalized assessment of specific „A (Amyloid) T (Tau) and N (Neurodegeneration)“ related microglial response rates. Tau pathology dominates the regional associations of these ATN biomarkers with microglial activation in primary and secondary tauopathies, while strong regional heterogeneity of ATN biomarker alterations needs to be considered at the individual patient level. Tau and Aβ related microglial response indices showed differential associations with cognitive performance and sTrem2-levels and may serve as a two-dimensional index for in vivo assessment of neuroinflammation in neurodegenerative diseases.

## Data Availability

The datasets used and/or analyzed during the current study are available from the corresponding author upon reasonable request. Supplementary information is available at MP's website.

