## Supplementary figures and images for "Individual regional associations between Aβ-, tau- and neurodegeneration (ATN) with microglial activation in patients with primary and secondary tauopathies"

### Supplemental Figure 1

**Supplemental figure 1**

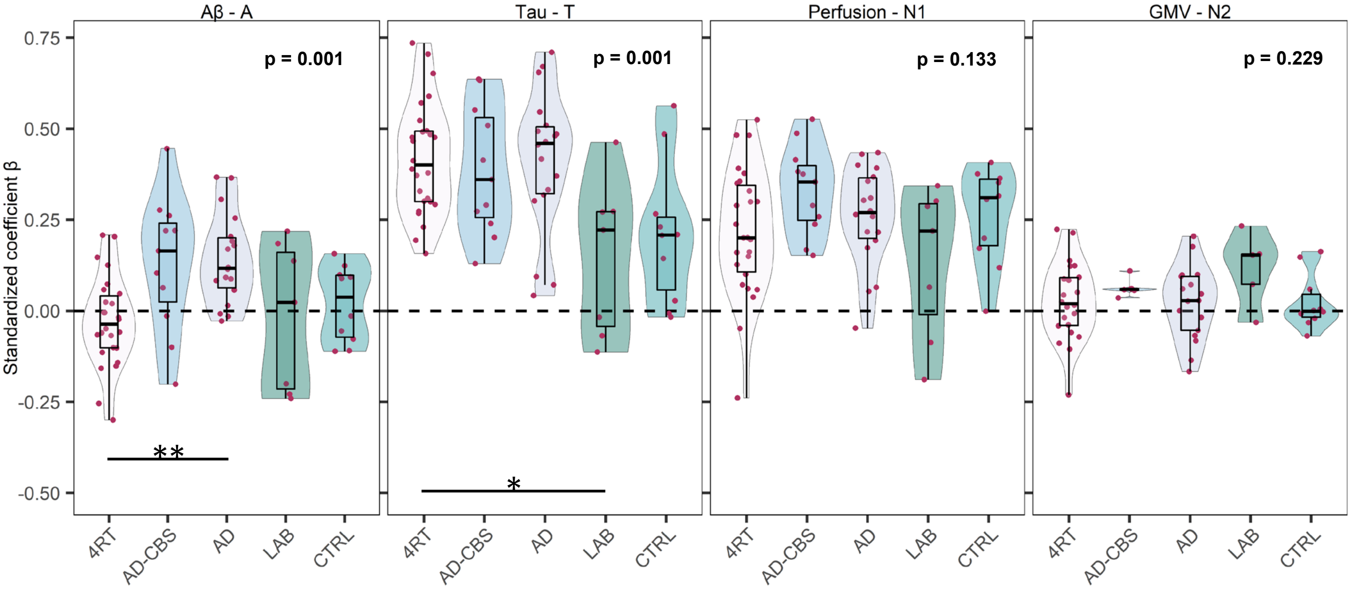

### Supplemental Figure 2

**Supplemental figure 2**

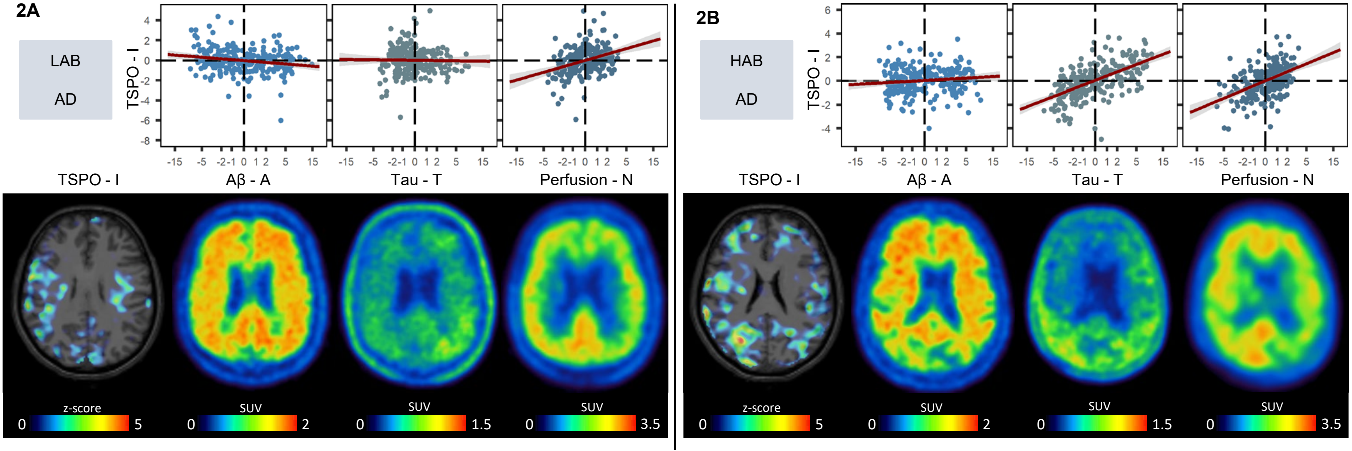
