## Supplemental Figure captions for "Individual regional associations between Aβ-, tau- and neurodegeneration (ATN) with microglial activation in patients with primary and secondary tauopathies"

**Supplemental Figure 1: Multiregional Regression Analysis to Determine the Association of ATN Biomarkers with Microglial Activation at the Individual Patient Level – Comparison Between Different Diseases**

A multiple linear regression model was performed (I = β_0_ + β_A_x_A_ + β_T_x_T_ + β_N1_x_N1_ + β_N2_x_N2_ + ε) at the single-patient-level and standardized coefficients (β) were derived for each ATN biomarker. Β-coefficients were visualized as a combined violin-box-plot and show the association of ATN biomarkers with microglia activation, each dot representing a single subject. Within each ATN biomarker group, the β-coefficients were tested for group differences between the diagnostic groups using ANOVA with age and sex as covariates. Multiple posthoc tests (dichotomous ANOVA, age and sex corrected) were performed if there was a significant group difference. Combined false discovery rate (FDR, ten p-values) and Bonferroni (four p-values) correction were applied to decrease risk of α-error-accumulation. * p < 0.05, ** p < 0.01, *** p < 0.001.

Abbreviations: AD, Alzheimer’s disease; AD-CBS, corticobasal syndrome with AD-pathology; 4RT, four-repeat tauopathies; LAB, low affinity binder; CTRL, healthy controls; TSPO, 18-kDa translocator protein; Aβ, β-amyloid; GMV, gray-matter-volume; ANOVA, analysis of variance; FDR, false discovery rate.

**Supplemental Figure 2: Specificity of Aβ and Tau Associated Microglia Activation**

All study participants underwent TSPO gene rs6971 SNP genotyping and classification into low-, medium- or high-affinity binding (LAB, MAB, HAB).

(**A**) Patient with LAB status and AD diagnosis. Partial regression of ATN-associated microglial activation within 246 brain regions. Brain sections of the respective PETs were shown with TSPO (z-score), Aβ (SUV), tau (SUV), and perfusion (SUV).

(**B**) Patient with HAB status and AD diagnosis. Same approach as (**A**). Note the strong regional association of Aβ and tau deposition with microglia activation which is absent in (**A**).

Abbreviations: AD, Alzheimer’s disease; TSPO, 18-kDa translocator protein; Aβ, β-amyloid; SUVr, standard-uptake-value-ratio; PET, positron-emissions-tomography.

,
