## Supplemental Table for "Individual regional associations between Aβ-, tau- and neurodegeneration (ATN) with microglial activation in patients with primary and secondary tauopathies"

**Supplemental Table 1: Demographics and Tracer Specific Control Cohorts**

Data are presented as mean ± standard deviation and number of frequency (n). Demographics were statistically tested by ANOVA or chi-square (χ²) test. With significant ANOVA group difference, multiple unpaired two-tailed Student's t-tests were performed.

Abbreviations: AD, Alzheimer’s disease; AD-CBS, corticobasal syndrome with AD-pathology; 4RT, four-repeat tauopathies; LAB, low affinity binder; MAB, medium affinity binder; HAB, high affinity binder; CTRL, healthy controls; TSPO, 18-kDa translocator protein; Aβ, β-amyloid; GMV, gray-matter-volume; ANOVA, analysis of variance; χ², chi-square; MoCA, Montreal-Cognitive-Assessment-Scale; MMSE, Mini-Mental State Examination; sTREM2, soluble triggering receptor expressed on myeloid cells 2.

| Supplemental table 1 | Neurodegenerative disease | | | | Controls | Tracer specific controls (z-score) | | | | | ANOVA / χ² |
| --- | --- | --- | --- | --- | --- | --- | --- | --- | --- | --- | --- |
|  | 4RT | AD-CBS | AD | LAB | CTRL | I  (TSPO) | A  (Aβ) | T  (late Tau) | N1  (early Tau) | N2  (GMV) | F  (p-value) |
| n | 26 | 11 | 18 | 7 | 10 | 13 | 13 | 14 | 14 | 13 |  |
| Age (years) | 70.8 ± 6.3 | 75.7 ± 5.5 | 70.7 ± 7.4 | 75.0 ± 6.5 | 69.5 ± 8.9 | 70.6 ± 7.5 | 70.6 ± 7.5 | 67.4 ± 9.5 | 67.4 ± 9.5 | 61.2 ± 8.4 | 3.254  (0.001) |
| Posthoc p-value Age \| 4RT \| AD-CBS \| AD \| LAB \| CTRL \| I \| A \| T \| N1; AD-CBS \| 0.082 \| \| \| \| \| \| \| \|; AD \| 0.958 \| 0.094 \| \| \| \| \| \| \|; LAB \| 0.210 \| 0.846 \| 0.217 \| \| \| \| \| \|; CTRL \| 0.641 \| 0.068 \| 0.690 \| 0.152 \| \| \| \| \|; I \| 0.930 \| 0.110 \| 0.970 \| 0.230 \| 0.733 \| \| \| \|; A \| 0.930 \| 0.110 \| 0.970 \| 0.230 \| 0.733 \| 1.000 \| \| \|; T \| 0.177 \| 0.008 \| 0.225 \| 0.035 \| 0.505 \| 0.277 \| 0.277 \| \|; N1 \| 0.177 \| 0.008 \| 0.225 \| 0.035 \| 0.505 \| 0.277 \| 0.277 \| 1.000 \|; N2 \| < 0.001 \| < 0.001 \| 0.001 \| < 0.001 \| 0.012 \| 0.002 \| 0.002 \| 0.042 \| 0.042 | | | | | | | | | | | |
| Sex | ♀ 13 / ♂ 13 | ♀ 7 / ♂ 4 | ♀ 9 / ♂ 9 | ♀ 2 / ♂ 5 | ♀ 7 / ♂ 3 | ♀ 7 / ♂ 6 | ♀ 7 / ♂ 6 | ♀ 9 / ♂ 5 | ♀ 9 / ♂ 5 | ♀ 8/ ♂ 5 | 4.829  (0.849) |
| MoCA | 22.8 ± 4.6 (n=24) | 17.6 ± 6.6 (n=10) | 22.4 ± 6.2 (n=14) | 22.0 ± 7.5 (n=3) | 28.6 ± 1.6 (n=9) | 26.6 ± 1.9 (n=13) | 26.6 ± 1.9 (n=13) | 25.2 ± 3.8 (n=5) | 25.2 ± 3.8 (n=5) | 23.8 ± 4.0 (n=13) | 5.010  (< 0.001) |
| Posthoc p-value MoCA \| 4RT \| AD-CBS \| AD \| LAB \| CTRL \| I \| A \| T \| N1; AD-CBS \| 0.002 \| \| \| \| \| \| \| \|; AD \| 0.806 \| 0.009 \| \| \| \| \| \| \|; LAB \| 0.769 \| 0.131 \| 0.878 \| \| \| \| \| \|; CTRL \| 0.001 \| < 0.001 \| 0.002 \| 0.027 \| \| \| \| \|; I \| 0.013 \| < 0.001 \| 0.015 \| 0.104 \| 0.311 \| \| \| \|; A \| 0.013 \| < 0.001 \| 0.015 \| 0.104 \| 0.311 \| 1.000 \| \| \|; T \| 0.268 \| 0.002 \| 0.229 \| 0.321 \| 0.174 \| 0.542 \| 0.542 \| \|; N1 \| 0.268 \| 0.002 \| 0.229 \| 0.321 \| 0.174 \| 0.542 \| 0.542 \| 1.000 \|; N2 \| 0.520 \| 0.001 \| 0.430 \| 0.531 \| 0.014 \| 0.102 \| 0.102 \| 0.538 \| 0.538 | | | | | | | | | | | |
